# Vaccine Hesitancy Among Adults in India: A Systematic Review and Meta-Analysis Protocol

**DOI:** 10.1101/2025.09.23.25336433

**Authors:** S Tejesh, A Y Nirupama, Varun Agiwal, Rovena Yazhini, Hotha Saisrinivas, Arijita Manna

## Abstract

**Background:** Vaccine hesitancy threatens public health progress, especially for populations lacking access to immunisation. Despite efforts to promote lifelong vaccination, adult immunisation is often overlooked. To create practical tools, understanding the level and types of vaccine hesitancy in adults is crucial. This review explores vaccine hesitancy in India’s adults, including psychological, social, and systemic factors influencing perceptions.

**Methods:** We will include observational studies and qualitative studies published in English that assessed vaccine hesitancy among Indian adults (above 18 years). We will search PubMed, Embase, Scopus, grey literature, and references. Two reviewers will independently review and screen studies and extract data using a standardised form. We will assess risk of bias using the JBI tools for observational studies and COREQ for qualitative studies. We will synthesise findings using either meta-analysis or narrative synthesis. sub-group analysis based on the type of vaccine and population. We will assess publication bias using funnel plots and Egger’s test.

**Results:** Since this is a protocol, results are not yet available. However, we expect to identify key trends related to the prevalence and determinants of vaccine hesitancy among Indian adults, particularly psychological, cultural, and systemic barriers. The synthesis should produce population-specific and vaccine-specific differences in vaccine hesitancy levels.

**Conclusion:** This review will synthesise evidence on adult vaccine hesitancy in India. The proposed evidence will be informing targeted interventions and will support policy-level efforts aimed at improving uptake of immunisations for adults and the vaccine confidence gap.

## Introduction

Vaccination is one of the most cost-efficient public health interventions (1), averting 3.5 to 5 million deaths per year from diphtheria, tetanus, pertussis (whooping cough), influenza and measles globally (2). Vaccine hesitancy, which is defined as the reluctance or refusal to vaccinate despite the availability of vaccines, is now a significant barrier to immunisation efforts (3). The World Health Organisation identified it as one of the top ten global health threats (4). In India, while there has been improvement in childhood vaccination, very few adults are vaccinated, and adult vaccination does not frequently feature in public health programmes (5,6).

Adults are at risk of the vast majority of diseases that can be prevented by vaccination because of declining immunity, absence of childhood vaccinations, age-related increases in susceptibility and comorbid chronic disease (7). Therefore, adults need to be vaccinated to protect themselves from diseases and promote healthy ageing. However, there are various challenges to adult vaccination, including lower awareness and education, sociocultural factors, community preferences, the lack of dedicated immunisation centre services for adult vaccination, and limited services for adult vaccination (7). A lack of strong recommendations from healthcare providers compounds the issues. Altogether, these barriers create a perfect storm, increasing the risk of developing illness and death from vaccine-preventable diseases in adults (7).

The burden of vaccine-preventable diseases (VPDs) in adults in India is substantial. A recently published meta-analysis by Satapathy et al. [2024] reported outcomes from 17 studies with 2,529 cervical cancer cases; 1,977 were HPV-positive, resulting in a pooled HPV prevalence of 85%, which underscores the relevance of HPV vaccination for adult women (8). Surveillance data from Kashmir observed that 27.5% of hospitalised adults with acute respiratory illness tested positive for influenza virus infections (9). In Pune, a measles outbreak occurred among vaccinated college students aged 18 to 24, illustrating the phenomenon of waning vaccine immunity (10). Systematic reviews indicate that Streptococcus pneumoniae is a leading cause of community-acquired pneumonia in adults in India (11). In addition, a hospital-based study from Kolkata submitted outcomes from 200 diphtheria cases. 75% of patients had a history of adequate immunisation; therefore, there may be gaps in booster dose immunisation (12).

Despite this burden of disease, adult vaccine utilisation in India is poor. A national survey of vaccination coverage among older adults estimated that only 1.5% of adults ever received the influenza vaccine, 0.6% for pneumococcal disease, 1.9% for typhoid and 1.9% for hepatitis B (13). Various studies have identified reasons for adult vaccine hesitancy, including low perceived need, safety concerns, lack of information and the absence of national guidelines (14–16). Adult vaccine hesitancy is high among health professionals and their families regarding zoster, pneumococcal, HPV, and influenza vaccines, primarily due to concerns about safety and limited information (16).

Although several individual studies have examined the prevalence and factors contributing to adult vaccine hesitancy in India, there has been no systematic attempt to review and collate this evidence. This study seeks to estimate the overall prevalence of adult vaccine hesitancy in India and describe the factors contributing to hesitancy by conducting a systematic review and meta-analysis. The study findings will help decision-makers with public health policy, help health care providers to manage vaccine-related queries, and assist in the development of targeted strategies to improve adult vaccine uptake in India.

### Review question

What is the prevalence, and what are the determinants of adult vaccine hesitancy in India?

### Methodology

#### Eligibility Criteria

##### Population

Studies including adults aged 18 and over in India from any situation or context, such as general population, healthcare workers or high-risk groups.

#### Study Types

This review will consider all observational studies (including cross-sectional, case-control, and cohort studies) examining the prevalence of vaccine hesitancy in adults in India, as well as qualitative studies exploring the determinants or barriers driving vaccine hesitancy. Only studies published in English will be included.

#### Outcome

##### Primary outcome

Prevalence of vaccine hesitancy among adults in India

##### Secondary outcomes

Determinants, barriers, or contributing factors of vaccine hesitancy among adults in India.

#### Information sources

A systematic search will be conducted in PubMed, Embase, and Scopus databases. In addition, reference lists of relevant studies will also be searched, and forward citation tracking will be done. To ensure full coverage, relevant national and international reports and grey literature found in Google searches will also be reviewed.

#### Search strategy

The search strategy will consist of a combination of medical subject headings (MeSH) and relevant free-text keywords, including core terms are: “adult vaccination,” “vaccine hesitancy,” “determinants,” “barriers,” “strategies,” “India”, “immunization,” and “vaccine uptake”. Boolean operators (AND, OR) will be used to ensure the terms are reduced and include the correct terms. The full search strategy will be adjusted for each database, and filters to restrict the studies to only those published in English will be added.

#### Selection process

All articles retrieved from the databases will be imported into Rayyan for screening. Once duplicates are removed, two independent reviewers (TS and AM) will screen the articles in two stages. In Level 1, titles and abstracts will be screened based on inclusion criteria, and in Level 2, full texts of selected articles will be assessed against the inclusion and exclusion criteria. Any discrepancies that arise between TS and AM will be resolved by way of discussion or by a third reviewer (HS). The complete selection process will be documented using a PRISMA flowchart.

#### Data collection process

Data from the studies included in this review will be extracted independently by TS and HS using a standardized data extraction form. Data extracted will include study characteristics (study design, setting, and methods), and participant characteristics (age and gender). The primary outcomes will relate to the prevalence of vaccine hesitancy and its determinant factors. Disagreements in the data extraction process will be resolved through discussion or by having a third reviewer (AM) involved. For any unclear data, study authors will be contacted; studies with authors that do not respond will be excluded.

#### Study risk of bias assessment

The risk of bias of the included studies will be determined according to the type of study design. The qualitative studies will be assessed following the COREQ (Consolidated Criteria for Reporting Qualitative Research) checklist. The quantitative studies, including observational studies, will be assessed using the Joanna Briggs Institute (JBI) critical appraisal tool or the Newcastle-Ottawa Scale. The quality assessment will be conducted by two independent reviewers, and any disagreement between reviewers will be resolved through discussion or through a third reviewer.

#### Effect measures

The effect measure for the primary outcome, the prevalence of adult vaccine hesitancy, will be reported as proportions (%) with 95% confidence intervals (CIs). For secondary outcomes for determinants of vaccine hesitancy, effect measures will be odds ratios (ORs) or risk ratios (RRs) at 95% CIs, which will be extracted as reported (if available, adjusted estimates will be prioritised). Qualitative outcomes will be summarised narratively.

#### Synthesis methods

Depending on the type and quality of included studies, we will synthesise the findings. If studies are quantitative and considered homogeneous, we will perform a meta-analysis to obtain pooled estimates of the prevalence of vaccine hesitancy. The analysis will be performed in RStudio (version 2025.05.1), and we will calculate pooled proportions with corresponding 95% confidence intervals. To determine heterogeneity, we will conduct the Chi^2^ test (with an alpha of 0.1) and measure heterogeneity using the I^2^ measure. If heterogeneity is low, then we will utilise the fixed-effect model; otherwise, we will use a random-effects model for the analysis. If there is too much heterogeneity for the data to be pooled, the results will be described in a narrative synthesis. If there are qualitative studies, we will do a meta-aggregation or thematic synthesis to identify common themes around the factor of vaccine hesitancy.

#### Subgroup analysis

Subgroup analyses will be conducted where data permit to examine variations in vaccine hesitancy based on important factors, including vaccine type, population type, and geographic context (e.g. urban versus rural). This will help to uncover patterns of hesitancy in relation to these distinct groups and geographic regions in India.

#### Reporting bias assessment

To evaluate the risk of reporting bias, publication bias will be assessed using funnel plots if there are at least 10 studies included. Additionally, Egger’s test will be applied to statistically assess small-study effects as necessary. For qualitative or narrative synthesis, selective reporting will be determined by comparing reported outcomes to outcomes listed in study protocols or trial registries where available.

## Results

Since this is a protocol, results are not yet available. However, we expect to identify key trends related to the prevalence and determinants of vaccine hesitancy among Indian adults, particularly psychological, cultural, and systemic barriers. The synthesis should produce population-specific and vaccine-specific differences in vaccine hesitancy levels.

## Discussion

This systematic review is anticipated to provide a strong evidence base regarding the prevalence and determinants of adult vaccine hesitancy in India. This review will provide a deeper understanding of the multidimensional barriers (individual, systematic) that influence intention to vaccinate, by systematically extracting and synthesizing information from both quantitative and qualitative studies. By providing subgroup analysis by vaccine type, population, and geographic context, we will be better able to target systemic areas needing intervention. Our findings will contribute to national immunisation policy, help healthcare providers address vaccine concerns, and steer localised awareness campaigns. This review will ultimately help strengthen evidence-informed strategies to promote adult vaccination uptake within India.

## Data Availability

All data produced in the present work are contained in the manuscript

## Declarations

### Availability of Data and Materials

The datasets generated and/or analysed during the current study will be available from the corresponding author upon reasonable request.

### Competing Interests

The authors declare that there are no competing interests, except for the fact that two team members are from the same field of study, which does not affect the objectivity of the research.

### Funding

This study did not receive any specific grant from funding agencies in the public, commercial, or not-for-profit sectors.

### Declaration of generative AI and AI-assisted technologies in the writing process

During the preparation of this work, the author(s) used Grammarly to refine the content. After using this tool/service, the author(s) reviewed and edited the content as needed and take full responsibility for the content of the publication.

## Notes

### Competing Interest Statement

The authors have declared no competing interest.

### Author Declarations

The study used ONLY openly available human data that were originally located at Google.

## References

1. Vaccination and Immunization Statistics [Internet]. UNICEF DATA. [cited 2025 Jul 4]. Available from: https://data.unicef.org/topic/child-health/immunization/

2. Vaccines and immunization [Internet]. [cited 2025 Jul 4]. Available from: https://www.who.int/health-topics/vaccines-and-immunization

3. Reticencia a la vacunación: Un desafío creciente para los programas de inmunización [Internet]. [cited 2025 Jul 4]. Available from: https://www.who.int/news/item/18-08-2015-vaccine-hesitancy-a-growing-challenge-for-immunization-programmes

4. Vaccine hesitancy is one of the greatest threats to global health – and the pandemic has made it worse [Internet]. [cited 2025 Jul 4]. Available from: https://www.gavi.org/vaccineswork/vaccine-hesitancy-one-greatest-threats-global-health-and-pandemic-has-made-it-worse

5. Immunization and Child Health | UNICEF India [Internet]. [cited 2025 Jul 4]. Available from: https://www.unicef.org/india/what-we-do/immunization

6. Lahariya C, Bhardwaj P. Adult vaccination in India: status and the way forward. Hum Vaccin Immunother. 16(7):1508–10.

7. Establishing Adult Vaccination Clinics in India: A Proposed Framework [Internet]. [cited 2025 Jul 4]. Available from: https://japi.org

8. Satapathy P, Khatib MN, Neyazi A, Qanawezi L, Said S, Gaidhane S, et al. Prevalence of human papilloma virus among cervical cancer patients in India: A systematic review and meta-analysis. Medicine (Baltimore). 2024 Aug 2;103(31):e38827.

9. Mir H, Haq I, Koul PA. Poor Vaccine Effectiveness against Influenza B-Related Severe Acute Respiratory Infection in a Temperate North Indian State (2019–2020): A Call for Further Data for Possible Vaccines with Closer Match. Vaccines. 2021 Oct;9(10):1094.

10. Bajaj S, Bobdey P, Singh N. Measles Outbreak in Adults: A Changing Epidemiological Pattern. Medical Journal of Dr DY Patil Vidyapeeth. 2017 Oct;10(5):447.

11. Ghia CJ, Dhar R, Koul PA, Rambhad G, Fletcher MA. Streptococcus pneumoniae as a Cause of Community-Acquired Pneumonia in Indian Adolescents and Adults: A Systematic Review and Meta-Analysis. Clin Med Insights Circ Respir Pulm Med. 2019 Jan 1;13:1179548419862790.

12. Outcomes of respiratory diphtheria in a tertiary referral infectious disease hospital - ProQuest [Internet]. [cited 2025 Jul 7]. Available from: https://www.proquest.com/openview/57e24dbdb643c5fec3717220996d7e67/1?pq-origsite=gscholar&cbl=46836

13. Rizvi AA, Singh A. Vaccination coverage among older adults: a population-based study in India. Bull World Health Organ. 2022 Jun 1;100(6):375–84.

14. Moosa MB, Josh D, Bobby R, Biju B, Sebastian J, John SB, et al. Factors influencing the hesitancy and refusal of vaccines in India: A study-using tool developed by WHO SAGE Working Group. Trends in Immunotherapy [Internet]. 2024 Apr 11 [cited 2025 Jul 7];8(1). Available from: https://ojs.ukscip.com/index.php/ti/article/view/479

15. Kataria I, Siddiqui M, Treiman K, Foley S, Anand M, Biswas S, et al. Awareness, perceptions, and choices of physicians pertaining to human papillomavirus (HPV) vaccination in India: A formative research study. Vaccine: X. 2022 Dec 1;12:100228.

16. Kalra N, Kalra T, Mishra S, Basu S, Bhatnagar N. Hesitancy for Adult Vaccines Among Healthcare Providers and their Family Members in Delhi, India: A Cross-Sectional Study. Dialogues in Health. 2022 Dec 1;1:100044.

